# Elucidating the Humoral Immune Response to SARS-CoV-2: Isolation and Characterization of Monoclonal Antibodies from Convalescent COVID-19 Patients

**DOI:** 10.1101/2025.08.01.25332666

**Authors:** Yiran Shen, Jason J. Thornton, Alexandria Voigt, Apichai Tuanyok, Cuong Q. Nguyen

## Abstract

The COVID-19 pandemic has underscored the importance of understanding the intricate mechanisms of the humoral immune response to SARS-CoV-2. This study aimed to elucidate the diversity and specificity of antibodies generated from convalescent COVID-19 patients by isolating and characterizing monoclonal antibodies (mAbs) targeting the SARS-CoV-2 spike protein. Employing cutting-edge technologies, including single-cell analysis and fluorescence-activated cell sorting, we successfully isolated live memory B cells secreting IgG antibodies from the peripheral blood of convalescent patients. A total of 17 mAbs were generated, encompassing various heavy and light variable genes, with only a few common between patients. In vitro assays demonstrated varying degrees of inhibition against wild-type and Omicron strains, highlighting discrepancies between ACE2 competition and actual neutralization capacity. Bio-layer interferometry and in silico docking analyses revealed unique binding motifs and mechanisms of action, with notable differences in neutralization abilities based on epitope specificity. Furthermore, animal experiments using K18-hACE2 transgenic mice demonstrated the therapeutic potential of these mAbs in preventing SARS-CoV-2 infection. This study provides novel insights into the humoral immune response to SARS-CoV-2 and highlights the importance of patient-derived mAbs as therapeutic agents for COVID-19 treatment and prevention.

## Introduction

B cells are essential components of the adaptive immune system, originating from the bone marrow and undergoing maturation to develop antigen specificity. These cells play a crucial role in the immune response to viral pathogens, including SARS-CoV-2, the virus responsible for COVID-19. B cells are activated upon encountering a viral antigen, and their role extends from recognizing and neutralizing viral pathogens through antibody production to establishing immune memory via memory B cells^1,2^. Memory B cells ensure rapid and robust responses upon re-exposure, providing long-lasting immunity^2^. B-cell response to these antigens, especially the S protein, is critical for understanding the body’s defense against COVID-19 and for vaccine development. Turner et al. reported that SARS-CoV-2-specific long-lived plasma cells (LLPCs) could persist for up to 11 months in the bone marrow, offering protection in mild COVID-19 cases^3^. Moreover, convalescent individuals exhibit convergent immunoglobulin heavy chain (IG_H_) specificities, particularly for the viral receptor-binding domain (RBD)^4^. Studies have demonstrated robust memory B cell responses in recovered individuals, maintained longer than serological IgG titers^5,6^, highlighting the potential of vaccines to induce strong B cell responses against evolving SARS-CoV-2 antigens^7^.

SARS-CoV-2’s ability to evade the immune system, including direct or indirect impacts on B cell function. The Omicron variant contains mutations like D614G and N501Y, which complicate the immune response^8–13^. These mutations enhance viral entry and fusion capabilities and challenge the efficacy of existing antibodies, aiding in immune evasion. Furthermore, SARS-CoV-2 infection has been associated with low levels of somatic mutations and defects in germinal center formation, potentially impairing the development of high-affinity memory B cells and plasma cells^14–16^.

Intensity and breadth of B-cell antiviral response reflected in monoclonal antibody production. Mechanisms of action of monoclonal antibodies for the treatment of viral infections include binding of neutralizing antibodies to the virions, opsonizing the virions or infected cells through binding for phagocytosis by phagocytosis, and promotion of death of the target cells through complement immobilization and activation of the membrane attack complex (MAC) or antibody-dependent cytotoxicity^1^. This research aims to elucidate the viral-specific B cell responses to SARS-CoV-2, focusing on identifying and analyzing specific monoclonal antibodies from convalescent patients and assessing their neutralizing properties through *in vitro* and *in vivo* assays. The structural interactions between antibodies and the spike protein of SARS-CoV-2 are examined to understand neutralization mechanisms and the effects of viral mutations. Animal experiments using K18-hACE2 transgenic mice were planned to validate the protective efficacy of these monoclonal antibodies. Through this analysis, this study seeks to enhance our understanding of B cell responses to SARS-CoV-2 and support the development of effective vaccines and therapies, contributing to global COVID-19 mitigation efforts.

## Materials and Methods

### Human samples

Peripheral blood mononuclear cells (PBMC) from post-convalescent COVID-19 donors were obtained from LifeSouth Community Blood Centers (Gainesville, FL). The volunteer donors had recovered from COVID-19 and were positive for SARS-CoV-2 antibodies at the time of blood donation. The donors had no prior clinically diagnosed autoimmune diseases. The samples were handled in a certified BSL2+ with Institutional Biosafety Committee-approved protocols.

### Ethics

This study received approval from the University of Florida’s Institutional Review Board, with the reference number IRB202001475. The recruitment of participants began on August 27, 2020, and is still ongoing. All participants provided their informed consent in written form by LifeSouth Community Blood Centers.

### Single-cell sorting

RBD (ACROBiosystems, Newark, DE) and S1 proteins (Novus Biologicals, Littleton, CO) were labeled with Dylight 488 and Dylight 650 (ThermoFisher Scientific, Waltham, MA), respectively, according to the manufacturer’s instructions. Convalescent patient PBMCs are stimulated as previously described^17^. Cells are stained with DAPI, anti-CD20 PE (Biolegend, San Diego, CA), and 1:1,000 dilutions of the labeled SARS-CoV-2 proteins. Live, CD20^+^ B cells reactive against both RBD and S1 were sorted into lysis buffer (provided in SMART-Seq HT Kit, Takara Bio Inc, Kusatsu, Shiga, Japan) using Sony SH800 Sorter (Sony, Minato City, Tokyo, Japan). We also employed Single-Cell Antibody Nanowells (SCAN, **Fig. S1**), which uses a dense array of nanowells fabricated of poly-dimethyl-siloxane (PDMS) containing individual cells, allowing for printing of corresponding molecules secreted by each cell^17^, with capture slides coated with anti-IgG (Invitrogen, Waltham, MA,) and anti-Ig (SouthernBiotech, Birmingham, AL). Captured slides were incubated with fluorescent-labeled RBD and S1 proteins to determine the B cell reactivity to these viral epitopes. Individual antibody-secreting B cells were manually picked using a micromanipulator and stored in lysis buffer as stated previously.

### Amplification and cloning of immunoglobulin genes

The variable (V) heavy (H) (V_H_DJ_H_) and light (L) (V_L_J_L_) chains of immunoglobulins (Igs) genes of individual cells were amplified using primer sets adapted from Rodda et al^18^ and Tiller et al^19^ for In-Fusion Cloning Kit (Takara Bio Inc, Kusatsu, Shiga, Japan). To express Igs, V_H_ and V_L_ were cloned into expression vectors pFUSEss-CHIg-hG1, pFUSE2ss-CLIg-hL2 and pFUSE2ss-CLIg-hK, respectively (Invivogen, San Diego, CA). The ligation products were transformed into competent Stellar™ Competent Cells (Takara Bio Inc, Kusatsu, Shiga, Japan,) and selected by Zeocin and Blasticidin resistance. Cloned plasmids were sequenced through GENEWIZ (Azenta, Chelmsford, MA) and aligned by NCBI IgBLAST to ensure sequences of chains were intact.

### Recombinant monoclonal antibody production

To produce recombinant monoclonal antibodies (mAbs), FreeStyle™ CHO-S cells (ThermoFisher Scientific, Waltham, MA, R80007) were transfected with corresponding H/L expression vector DNA through FectoCHO® Expression System (PolyPlus, Vectura, France) as instructed by the manufacturer. On day 7 post-transfection, the cell culture was centrifuged and purified through EconoFit UNOsphere SUPrA on NGC™ Chromatography Systems according to the manufacturer (Bio-Rad, Hercules, CA). Protein concentration was quantified by Bradford protein assay (Quick Start™ Bradford 1x Dye Reagent, Bio-Rad, Hercules, CA) and the integrity of the Ig fractions with joined H/L chains was determined by Mini-PROTEAN TGX Stain-Free Precast Gels (Bio-Rad, Hercules, CA) by sodium dodecyl sulfate–polyacrylamide gel electrophoresis (SDS-PAGE).

### Determination of antibody binding activity to SARS-CoV-2 RBD

The anti-SARS-CoV-2 neutralizing antibody titer serological assay kit (ACROBiosystems, Newark, DE) was used to measure the binding capacity of mAbs, according to the manufacturer. The samples were diluted to 2ug/mL, 1ug/mL, and 0.5ug/ml. The samples were added to wells containing 0.3μg/mL of HRP-conjugated RBD that binds to the immobilized human ACE2 protein pre-coated on 96-well plates. After adding the substrate and stop solutions, the final absorbance was detected at 450 nm. The results of the detected signal were normalized to the percentage of inhibition compared to the positive control, and a percentage of inhibition of 20% was considered inhibitory. CR3022 (BEI Resources, NIAID, NIH: Monoclonal Anti-SARS Coronavirus Recombinant Human Antibody, Clone CR3022 (produced in HEK293 Cells), NR-52481) was used as a neutralizing antibody positive control.

### Validation of antibody-neutralizing abilities using pseudovirus

Ha-CoV-2 particles from Virongy Biosciences, a hybrid SARS-CoV-2 virus-like particle (VLP) encapsulating an alphavirus-derived RNA genome for rapid luciferase expression, with SARS-CoV-2 structural proteins (S, M, N, and E) on its surface. The mAbs were prepared at a concentration of 50 µg/mL from the stock solution, and a 5-fold serial dilution of each mAb was performed using Dulbecco’s Phosphate-Buffered Saline (DPBS) (Corning Incorporated, Corning, NY). Ha-CoV-2 particles [Omicron (B.1.1.529)] are pre-incubated with the serially diluted mAbs for 30 minutes at room temperature. Following the ready-to-use HEK293T(ACE2/TMPRSS2), cells were seeded in a 96-well plate, the pre-incubated mixtures of Ha-CoV-2 particles and mAbs were added to the seeded cells, and the infection process took place over 18 hours. Cells were lysed post-infection. A luciferase assay was performed using reagents provided with the kit and following the manufacturer’s instructions. Anti-SARS-CoV-2 RBD AM180 (Acro Biosystem, Newark, DE) and Adintrevimab (MedChemExpress, Monmouth Junction, NJ) were used as positive controls.

### Binding kinetics and affinity assessment of mAbs by Bio-Layer Interferometry (BLI)

The binding kinetics and affinity of mAbs to the SARS-CoV-2 S protein were evaluated by BLI using the Gator label-free bioanalysis system (GatorBio, Palo Alto, CA). The anti-His biosensor (GatorBio, Palo Alto, CA) was pre-equilibrated with PBS before immobilization with 7.5 μg/ml SARS-CoV-2 S protein (Super stable trimer) (Acro Biosystem, Newark, DE) and SARS-CoV-2 spike trimer (BA.1.1/Omicron) (Acro Biosystem, Newark, DE). All steps were performed under default temperature at 400 rpm for antigen loading and 1,000 rpm for the remaining steps. All reagents were prepared in PBS. S proteins (7.5 μg/ml) were immobilized on the anti-His biosensor at a level of ∼1 nm. After a 60-second baseline step in PBS, the antigen-loaded biosensors were exposed (180 seconds) to analytes (mAbs) (ranging from 16.7 nM to 0.4 nM, in 3-fold serial dilutions) to measure binding, and then the sensor was immersed in PBS (180 seconds) to measure dissociation of the analyte from the surface of the antigen-loaded biosensor. Further analysis was performed using GatorBio analysis software to align the baseline prior to the binding phase reaction, reference subtraction (to the blank sensor), inter-step corrections (to the binding step), and fitting single- or dual-phase binding and dissociation kinetics to determine the k_on_, k_off_, and K_D_ (binding constants based on the ratio of the individual rate constants). Anti-SARS-CoV-2 RBD AM180 (Acro Biosystem, Newark, DE) and Adintrevimab (MedChemExpress, Monmouth Junction, NJ) were used as positive controls.

### Antibody epitope prediction

AbEMap from ClusPro (https://abemap.cluspro.org/cobemap/index.php) was used for antibody-based epitope mapping. The Protein Data Bank (PDB) files generated by ABodyBuilder2 serve (https://opig.stats.ox.ac.uk/webapps/sabdab-sabpred/sabpred/abodybuilder2/) as inputs for AbEMap. The prefusion 2019-nCoV spike glycoprotein with a single receptor-binding domain up (PDB ID: 6VSB) is selected as the antigen for docking to identify the antigen residues at the interface of the antibody-antigen binding.

### SARS-CoV-2 Propagation

Vero E6 cells (African Green Monkey Kidney Epithelial Cells expressing high endogenous Angiotensin-Converting Enzyme 2, BEI Resources, NIAID, NIH: NR-53726) to a 75-80% confluent monolayer were grown in 75 cm2 flasks (T-75). The cells were cultured in Minimum Essential Medium (MEM) Eagle (EMEM) supplemented with 10% Fetal Bovine Serum (FBS) and incubated at 37°C in a 5% CO2 atmosphere. Once the cells reached confluency, the flasks were transferred to BSL-3 lab. Frozen stocks of SARS-CoV-2 WA1/2020 culture (BEI Resources, NIAID, NIH: NR-52281) and B.1.1.529 culture (BEI Resources, NIAID, NIH: NR-56496) were thawed at 37°C. The culture medium was aspirated from the T-75 flasks, and the thawed viral culture was then inoculated onto the Vero E6 cell monolayer. Following a 1-hour incubation period, 12 mL of freshly prepared infection medium (EMEM with 2% FBS) was added to the flask, which was then incubated under the same conditions as previously stated. The Vero E6 cell monolayer was checked daily for cytopathic effect (CPE) signs. Viruses were harvested 3 days post-infection for WA1 and 5 days post-infection for B.1.1.529 from the culture medium. If sufficient CPE was not observed, the supernatant of the T-75 viral culture was collected and then infected with freshly prepared Vero E6 to obtain a higher viral yield. Once ready for harvest, the culture medium containing cells and viruses was collected in a 15 mL conical tube and centrifuged at maximum speed for 5 minutes to pellet the cells. The supernatant (viral specimen) was then collected, aliquoted into 500 µL volumes in Nalgene cryovials, labeled, and stored in a −80°C freezer for future use.

### Propagation and determining the plaque-forming units of SARS-CoV-2 virus

A T-75 flask containing a 75-80% confluent monolayer of Vero E6 cells, grown in EMEM supplemented with 10% FBS, was counted in a BSL-2 laboratory. 2.5 × 10^5^ cells per well were seeded in triplicate in 12-well plates (Corning Incorporated, Corning, NY) and incubated overnight at 37°C with infection medium. A 10-fold serial dilution of the viral stock, ranging from 10 to 10^6^, was prepared using an infection medium in 2 mL screw-cap tubes. The cell culture medium was removed from the 12-well plates, and 200 µL of the diluted viral solution was added to each well of the cell monolayer. A well with infection medium alone was included in each replicate as a negative control. The plates were gently rocked in left-to-right and front-to-back motions to facilitate the absorption of the viral solution by the monolayer. The plates were incubated at 37°C with 5% CO_2_ for 1 hour, rocking them every 15 minutes. To determine the plaque-forming units, equal amounts (1:1) of Gibco™ MEM (2x) (ThermoFisher Scientific, Waltham, MA) and 3% Carboxymethylcellulose (CMC) (Sigma-Aldrich, St. Louis, MO) were mixed to make Liquid Overlay Medium (L-OM). After the 1-hour incubation, 1.5 mL of L-OM was added to each well. The plates were incubated at 37°C with 5% CO2 for an additional 3 days. At the end of the 3-day incubation, 10% formaldehyde solution was added directly to the L-OM and incubated in the plate at room temperature for 1 hour for fixation. The cells were gently washed once with sterile PBS. 0.5% crystal violet solution was added to each well and incubated at room temperature for 15 minutes. Cells were washed twice with distilled tap water and blot-dried on paper towels. The uniform monolayer in the negative control was used as a reference to count the plaques, which appeared as clear circles on a purple monolayer of cells. The number of plaques observed per well was recorded at each viral dilution to calculate plaque-forming units (PFU).

### Plaque-reduction neutralization test (PRNT) for SARS-CoV-2 Infection

Monoclonal antibodies were initially prepared at a concentration of 30 µg/mL from the stock solution. A 3-fold serial dilution of each mAb was performed using an infection medium. The viral stock was diluted to achieve an approximate concentration of 50 PFU per well. For each well to be tested, an equal volume of mAbs and virus was mixed, resulting in a starting concentration of 15 µg/mL for the mAbs in the mixture. The assays were performed in duplicate and included a well with the virus alone in each replicate as a positive control.

### Administration of mAbs in SARS-CoV-2-infected K18-hACE2 mice

The homozygous K18-hACE2 mice were bred at the University of Florida (Breeding Project ID #3276). Mating pairs of hemizygous K18-hACE2 mice were purchased from Jackson Laboratory (034860–B6.Cg-Tg(K18-ACE2)2Prlmn/J). F1 mice were anesthetized for ear punching for DNA extraction and subjected to zygosity analysis. The F1 zygosity (homozygosity) was confirmed via an SNP assay (The Jackson Laboratory’s Protocol 38275: Sanger sequencing assay – Chr2_rs13476660-SEQ) by Transnetyx Inc. This assay detects an SNP between C57BL6/J and SJL/J (homozygous mutant), located approximately 4.7 kb from the transgene integration site. For infection, mice were intranasally inoculated with 20 µL of 860 PFU SARS-CoV-2 inoculum drop-by-drop into both nostrils until fully inhaled, and the control group was inoculated with an equivalent volume of PBS. For the test groups, each mouse received a single intraperitoneal injection of a 20 ug dose of single/combined antibodies 48 hrs before infection. Mouse body weights are monitored and recorded for 14 days, and dead mice are removed from the weight monitoring.

### Histological examination

Necropsies were performed on all animals following humane euthanasia by authorized personnel, and the principal investigator or an authorized representative performed diagnostic necropsies. The study investigator determined the tissue sampled for histopathologic examination (lung). All collected tissues were immersion-fixed in 4% paraformaldehyde (PFA) in PBS. Each set of PFA-fixed tissue samples was assigned a unique pathology accession number using the University of Florida Animal Care and Use Program’s Standard Operating Procedure. This accession number was used to track the tissues within the laboratory. All tissues were processed in the University of Florida College of Veterinary Medicine’s histology laboratory for this study. The tissue samples were trimmed, routinely processed with the tissue processor, and embedded in paraffin wax. Sections of the paraffin-embedded tissues were trimmed five to six μm thick for histology and mounted on pre-labeled, unstained positively charged glass slides using the ThermoScientific SlideMate Slide Printer. The histology slides were deparaffinized, stained with hematoxylin and eosin (H&E) stain, and cover slipped. The slides were scanned and digitized at 400X using the Leica Aperio Versa 200.

### Statistics Analysis

Statistical analyses were performed using one-way ANOVA and Mann-Whitney test where indicated (Prism 8, GraphPad, La Jolla, CA). P values ≤ 0.05 were considered significant.

## Results

### Generation of SARS-CoV-2-specific antibodies

The rapid antigenic drift of SARS-CoV-2 mediated by the spike (S) protein is exploited for host cell entry via the angiotensin-converting enzyme 2 (ACE2) receptor. B cells elicit an adaptive immune response by producing spike protein-specific antibodies that effectively bind to and neutralize the virus. The high affinity of these antibodies for the spike protein results from a precisely matched antigen-antibody interaction by the heavy and light chains of the antibodies. Nevertheless, the dynamic nature of SARS-CoV-2 is characterized by frequent mutations in the spike protein, particularly in regions encoding the RBD. These mutations lead to changes in the antigenic epitopes of the virus, thereby facilitating escape from recognition and neutralization by existing antibodies, compromising the efficacy of the humoral immune response. As a result, we sought to generate and determine the functionality of SARS-CoV-2-specific antibodies from memory B cells of convalescent patients, particularly antibodies reactive against RBD and S1 mutants. Using the SCAN method and FACS, we identified and isolated live memory B cells (Calcein^+^CD27^+^CD20^+^) secreting IgG antibodies specific to the RBD. We first identified B cells specific to the Wuhan-Hu-1 S1 and RBD in patient 1 (**Fig. 1A, C**). To determine if we could identify and isolate B cells with specific mutations, we tested B cells against S1 protein of Wuhan, Delta, and D614 using SCAN, in which P1 was positive for Delta and D614 and P2 for Wuhan, Delta, and D614 with IgG isotype (**Fig. 1B**). Using flow cytometry, we could identify memory B cells positive for S1 and RDB (0.71%) in patient 1 (**Fig. 1C**). Profiling B cells of patient 2, we were able to identify B cells reactive to multiple S1 mutations, including N501Y/Delta S, N501Y/D614G, and D614G^/^N501Y^/^Delta S (**Fig. 1D**).

**Figure 1:**
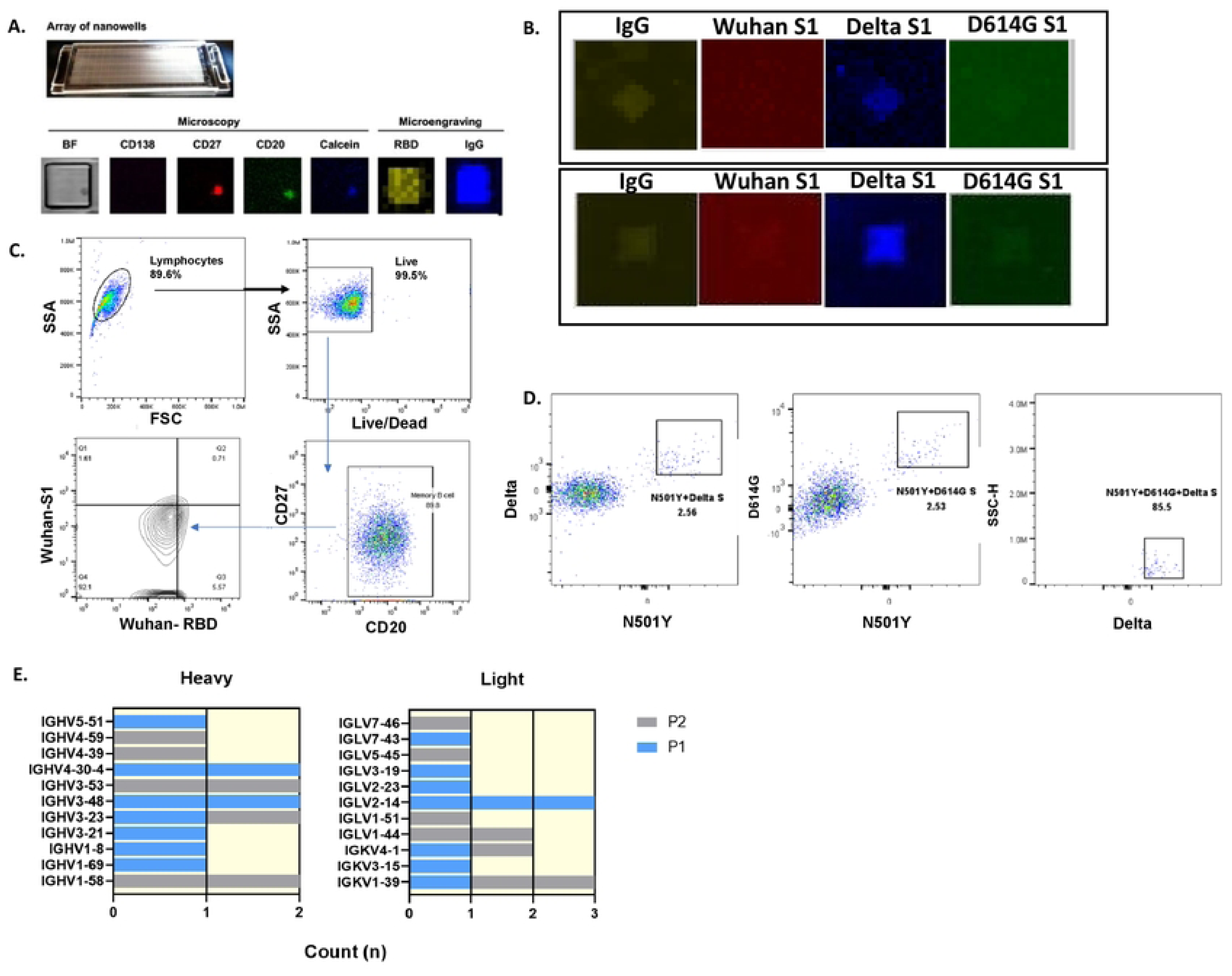
Identification and isolation of live memory B cells against SARS-CoV-2 from convalescent plasma. The representative image of SCAN for identified live memory B cells (Calcein+CD138^-^CD27+CD20+) secreting IgG antibodies specific to the **A**) SARS-CoV-2 RBD and **B**) Wuhan S1, Delta S1, and D614G S1 of patient 1 (top) and patient 2 (bottom). **C**) FACS gating strategy for live memory B cells against either Wuhan S protein or RBD from CD19- enriched B cells for patient 1. **D**) Gating live memory B cells against the S protein of variant (single-, double- or triple-positive) from CD19-enriched B cells for patient 2. **E**) Profile of 11 distinct heavy variable genes and 11 distinct light variable genes of patient 1 (P1) and patient (P2).

Using a micromanipulator, we picked 18 B cells that were Delta^+^/D614G^+^ from nanowells of patient 1. We sorted 15 B cells, there were D614G^+/^N501Y^+/^Delta^+^ via FACS of patient 2. We generated 17 mAbs, encompassing 11 heavy variable genes and 11 light variable genes (**Fig. 1E)**. Notably, significant variability was observed in the expression of heavy-chain and light-chain genes across these two subjects. Among the 22 variable genes analyzed from both heavy and light chains, only IGHV3-23, IGHV1-58, and IGKV1-39 were common between patients 1 and 2. Each individual exhibited variations in gene expansion, with certain genes being more prevalent. For instance, IGHV4-30-4 and IGHV3-48 were found twice for patient 1, IGHV3-53 was found twice for patient 2, and IGLV2-14 was found three times in patient 1, with IGLV1-44 appearing twice, compared to other genes, which were present only once. In summary, using SCAN and FACS, we have identified shared and unique B cell receptor clonotypes that were specific to various combinations of mutations of SARS-CoV-2 in convalescent patients.

### Monoclonal SARS-CoV-2 antibodies exhibited differential binding and inhibitory effects

Heavy and light chains of individual B cells described in **Fig. 1E** of patients 1 and 2 were expressed, and monoclonal antibodies (mAbs) were isolated as described in the methods (**Table 1**). To characterize these mAbs for their neutralizing activity, we first examined their inhibitory capacity against the wild-type strain that was selected for and, more importantly, determined their cross-reactivity against the emerging Omicron strains using a competitive ELISA, wherein the human ACE2 receptor is pre-affixed to test wells. We tested nine isolated mAbs with CR3022 (a commercial monoclonal antibody against SARS-CoV) as a positive control. As presented in **Fig. 2A**, the nine mAbs demonstrated varying degrees of inhibition, ranging from approximately 10% to 50%. Against the wild-type SARS-CoV-2, most mAbs tested showed higher inhibition at higher concentrations, notably for P1-A10 and P1-B5. Against Omicron, all nine mAbs showed similar effects at the three concentrations; overall, they showed better inhibition than against the wild type. except P1-C9. To further substantiate these findings, a pseudoviral infection assay was employed to measure the real-time efficacy of mAbs in preventing viral infections. This assay, utilizing pseudovirus particles Ha-CoV-2 that contain SARS-CoV-2’s complete structural protein set rather than the S protein alone, measures viral uptake into HEK293T cells expressing ACE2 and TMPRSS2. Six antibodies from P1, showing higher neutralization potential in the competitive ELISA, and eight antibodies from P2, identified using several S proteins with specific point mutations, were examined alongside two controls (Adeintervimab and AM180) differing in variant blocking efficacy (**Fig. 3B**). The real-time blocking experiments indicated that P1 mAbs, despite their broad RBD-binding capabilities using the ELISA, generally failed to prevent Omicron infections, with some even enhancing viral uptake as indicated by inhibition percentages exceeding 100%. This suggests a lack of correlation between ACE2 competition and neutralization activity. Interestingly, CN12, CN13, CN16, 8+14 from P2, and control AM180 similarly facilitated viral uptake. Still, most P2 mAbs effectively blocked virion uptake with a blockage of less than 50% at various concentrations, with 7+11 outperforming others across concentrations as low as 2 ug/mL, comparable to control Adeintervimab and CN8 at 2 ug/mL. This highlights that selecting memory B cells with multiple S proteins featuring single mutations facilitates the identification of antibodies targeting specific variant mutations.

**Figure 2:**
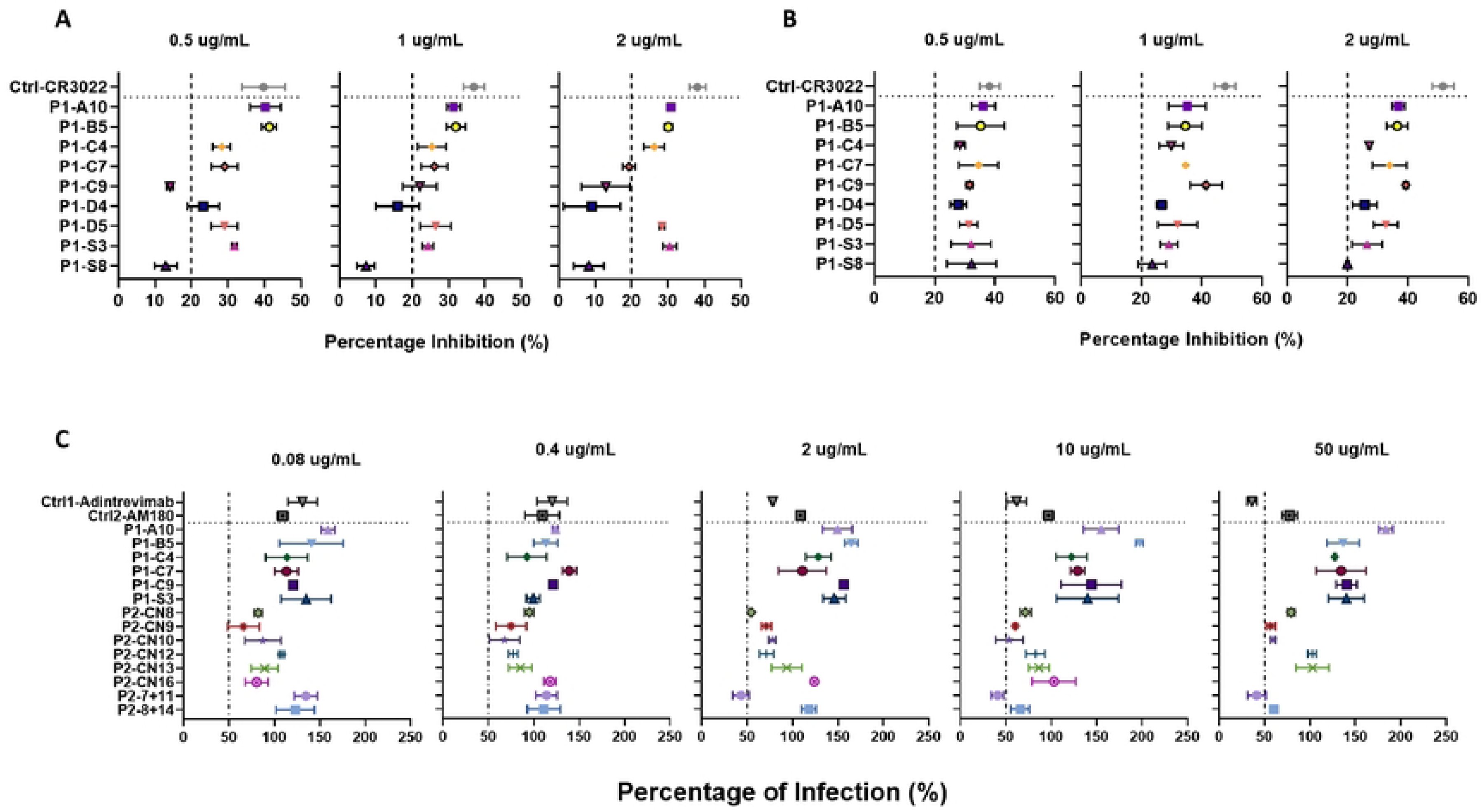
Summary of *in vitro* assay for the ability to inhibit SARS-CoV-2 infection. Competitive enzyme-linked immunosorbent assay to determine whether mAbs produced by patient 1 can inhibit (**A**) wild-type or Wuhan (**B**) Omicron SARS-CoV-2 RBD from binding to the ACE2 receptor. Three mAb concentrations (2 µg/mL, 1 µg/mL, and 0.5 µg/mL) were tested. mAbs were listed on the Y-axis, and the mean ± SEM percent inhibition was plotted on the X-axis. 20% is the threshold for positivity as supplied by the manufacturer. **C**) Pseudovirus neutralization assays for mAbs produced by patients P1 and P2. The ability of mAbs from P1 and P2 to block Omicron SARS-CoV-2 infection was tested across different concentration ranges, with the concentrations used in the experiments listed separately. Using a luciferase-based detection method, the infection rate was calculated based on the number of pseudovirus particles containing fluorescent markers within the cells. The results are plotted on the X-axis as mean ± SEM, with the Y-axis indicating the samples used. The threshold for positivity is 50%.

**Figure 3:**
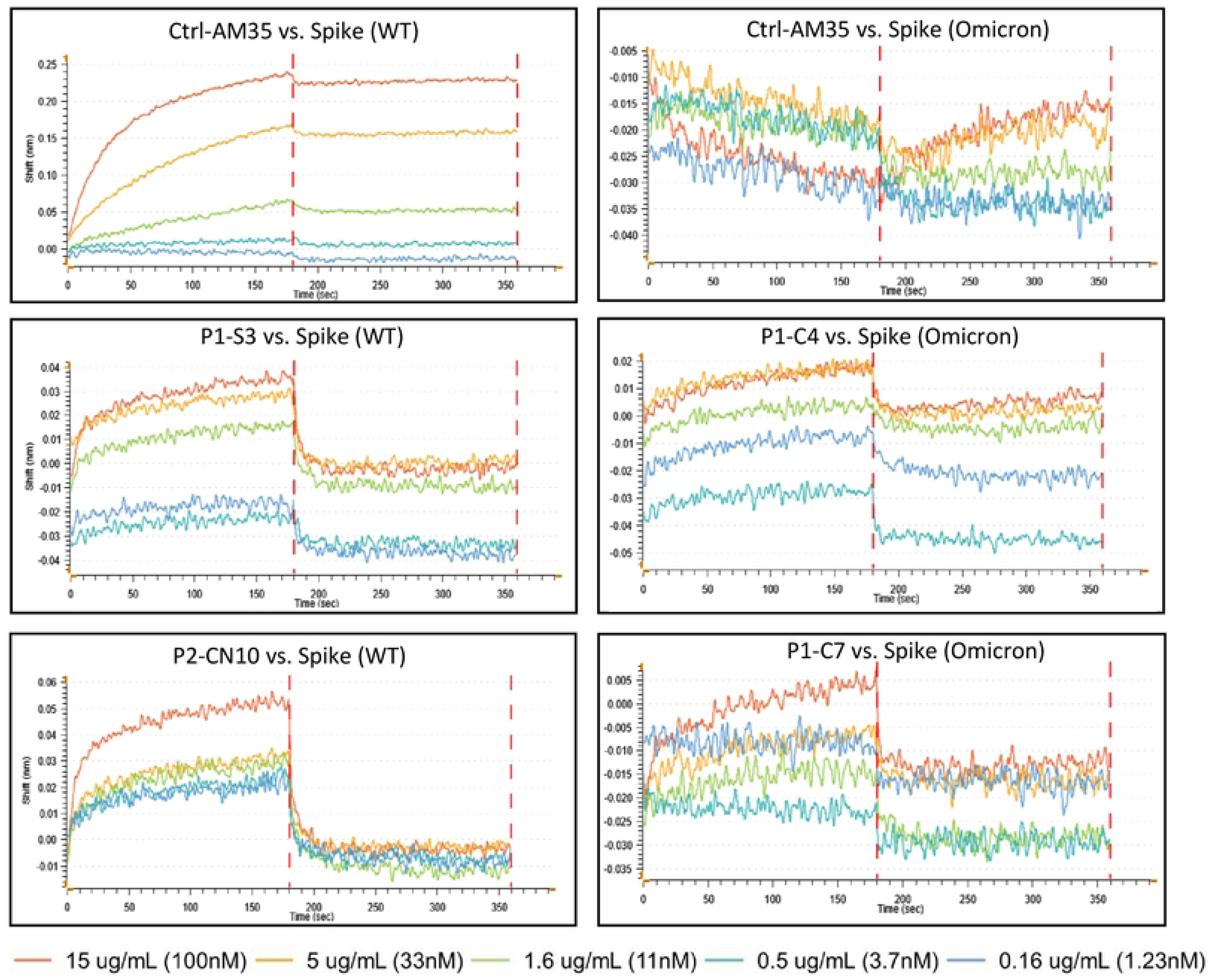
Representative images of kinetic analysis of selective monoclonal antibodies (mAbs) binding to the S protein at different concentrations. The binding kinetics of each mAb to the wild-type (WT) S protein (left) and the Omicron S protein (right) in the bound state and dissociated state (Shift) were detected using bio-layer interferometry (BLI). Different colors represent different concentrations, ranging from 1.23 nM to 100 nM. The Y-axis represents normalized signal shift, and the X-axis shows the time course from binding (first dashed line) to dissociation (second dashed line).

Lastly, antibody binding kinetics are critical to understanding antibody characterization and are often quantified by dissociation constants, thereby measuring affinity by the dynamic binding and dissociation events in a buffer system. We employed BLI assay to evaluate the K_d_ of mAbs from both P1 and P2 against the S protein from wild-type and Omicron strains (**Fig. 3**). AM35 was used as a control and for AM180 due to its diminished neutralizing ability, which was observed in the Omicron pseudoviral infection assay. The findings were unexpected. Although certain monoclonal antibodies (such as P1-S3 and P2-CN10 against the wild-type virus, and P1- C4 and P1-C7 against the Omicron variant) exhibit patterns similar to the positive control (AM35) during binding and dissociation (**Fig. 3**), i.e., as the test concentration levels increase (from 1.23 nM to 100 nM), binding shows an increase in shift (nm), while dissociation shows a more rapid decrease in shift (nm). All tested mAbs performed near or below background levels compared to control mAbs; it is worth noting that AM35 also showed poor results in binding tests with the Omicron spike protein. indicating that the binding assay alone may not adequately reflect the true inhibitory potential of patient-derived mAbs.

### Antibodies bind to different epitopes of SARS-CoV-2 structural proteins

The data indicated that the neutralizing capacity and binding kinetics of the mAbs were different. To further determine the precise epitopes that mAbs were physically interacting with, we performed structural analysis to reveal the binding motifs of the mAbs to SARS-CoV-2 structural proteins. Using data from the CoV-AbDab database (last updated: 13th June 2023), the V genes for heavy and light chains of published and patented antibodies against SARS-CoV-2 were plotted against the different CDR3 lengths (**Fig. 4**). The density of color in the plot represents the overall count of corresponding antibodies in the database. The V genes of our mAbs were identified with a green frame in the figure. The analysis revealed that our mAbs were diverse, with some of their V genes having a higher incidence in the overall count in the H/L chain. For instance, IGHV3-53, with a CDR3 length equal to 11, and IGKV1-39, with a CDR3 length equal to 9 and 10, separately, had counts that were in the highest range (**Fig. 4A**). To further analyze the sequence properties of the H/L chains, the combination of VJ genes from the database was plotted in the same way as presented in **Fig. 4B**. However, none of our generated mAbs showed an identical profile to the published and patented antibodies in the database.

**Figure 4:**
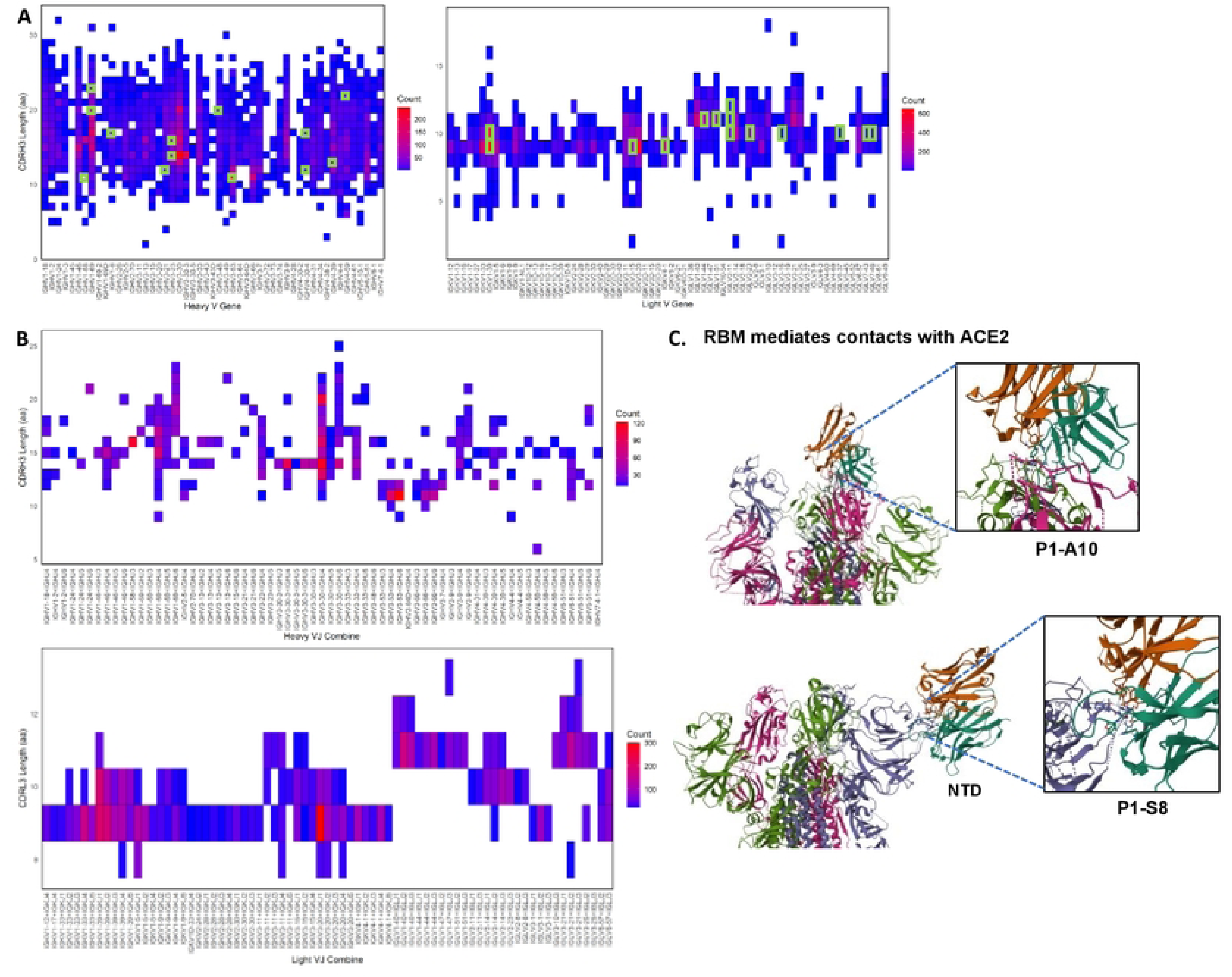
Profile of published/patented antibodies against SARS-CoV-2 with comparison to lab-produced mAbs from P1 and P2. **A**) Distribution and counts of heavy (left) and Light (right) variable genes according to CDR3 length. Variable genes with the same genes and CDR3 length as our lab-produced mAbs were labeled (green). **B**) Distribution and counts of heavy (up) and light (down) variable plus joining (VJ) genes according to CDR3 length. **C**) Differences in the location and mode of binding of antibodies to antigens according to *in silico* docking. RBM: Receptor-binding motif, ACE2: Angiotensin-converting enzyme 2, NTD: N-terminal domain of spike protein.

Due to the unique characteristics of our mAbs, an *in silico* docking analysis was performed using ClusPro 2.0 to reveal the differences in location and mode of binding of these antibodies to SARS-CoV-2 structural proteins (**Fig. 4C**). A reference antibody (Etesevimab by Eli Lilly) was used to validate the accuracy of structural docking, and the results were similar to the published data, predicting that the CDR regions of Etesevimab bind to the RBD regions on the spike trimer. The same selection criteria were applied to our two mAbs, P1-A10 and P1-S3, which were derived from memory B cells reactive against S1 protein and RBD. P1-A10 demonstrated the highest inhibition ability against the wild-type RBD, while P1-S3 showed the lowest in the previous competitive ELISA experiments. The docking analysis predicted that P1-A10 binds to the receptor-binding motif (RBM) in the RBD region that mediates contacts with ACE2, which is a highly immunogenic region and a key target for neutralizing antibodies and vaccine development, whereas P1-S3 binds to the N-terminal domain (NTD) in the S1 region, where the epitopes may not be easily accessible for antibody binding. Partly explaining the differences in neutralization ability in previous experiments.

### Reactivity of monoclonal antibodies against SARS-CoV-2 infection *in vitro*

The S protein of SARS-CoV-2 undergoes several conformational changes during the process of viral infection, which are crucial for the virus to bind to ACE2 and facilitate the fusion of the viral and host cell membranes. The RBD within the S1 subunit is initially in a “down” conformation, rendering it inaccessible for binding to the ACE2 receptor. To initiate infection, at least one RBD in the S protein trimer must switch from the “down” to the “up” conformation, exposing the RBD and making it accessible for binding to the ACE2 receptor on the host cell surface. During infection, transitioning from “down” to “up” conformation is a dynamic process, with the RBDs in the S protein trimer switching between these two states, providing a higher chance for antibody accessibility. To test the effectiveness of mAbs in preventing infection, we conducted the PRNT. As illustrated in **Fig. 5**, nine mAbs were selected, as they have been previously tested for their inhibition abilities against the original strain. Compared to the control antibody Adintrevimab, which demonstrated a higher neutralizing ability reflected by a significantly reduced plaque number, antibodies P1-A10 and P1-C7 showed consistent inhibition throughout the selected concentration range (from 31.25 ng/mL to 2000 ng/mL). Interestingly, P1-C9 exhibited the strongest neutralizing ability, almost comparable to the Adintrevimab positive control at the lowest concentration (31.25 ng/mL) and P1-A10 at 500 ng/mL. This suggests that the ability to bind to RBD is guided in selecting high-affinity antibodies against infection.

**Figure 5:**
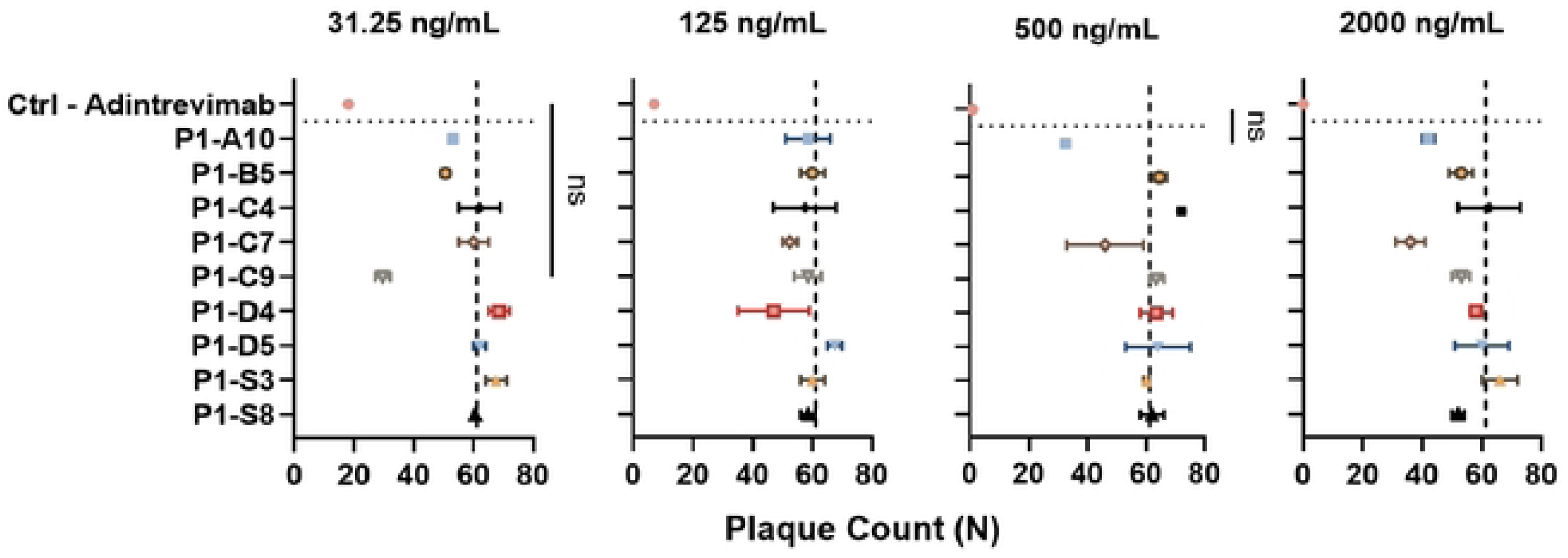
PRNT assays demonstrated the potential inhibitory ability of nine mAbs against SARS-CoV-2 infection at different concentrations (31.25 ng/mL to 2000 ng/mL). Viral inhibition was measured by plaque counts (N). The number of plaques is opposite to the inhibitory capacity. The dashed line on the X-axis indicates the mean counts of the plaque from all samples’ negative controls as a reference. Ns: not statistically significant. Statistical analysis was performed using one-way ANOVA.

### The therapeutic effect of monoclonal antibodies against SAR-CoV-2 infection in the mouse model

To evaluate the *in vivo* efficacy of mAbs against SARS-CoV-2, we evaluated their therapeutic potential in mice infected with SARS-CoV-2 to determine their ability to provide protection against viral challenge and mitigate associated lung pathological outcomes. Infected mice treated with P1-A10 were completely protected 14 dpi. P1-C4 provided 100% protection but reduced to 75% at 14 dpi. P1-S3 showed 50% survival at 9 dpi in comparison to 30% of infected without prophylactic treatment died at 6 dpi (**Fig. 6A**). Infected mice treated with the mAbs showed no weight loss at 6dpi, whereas the infected mice with no mAb treatment showed a slight reduction in weight (**Fig. 6B).** Examining the lung pathology, the treatment with mAbs significantly decreased pulmonary inflammation associated with SARS-CoV-2 infection in mice (**Table 2**). Except for C4, pneumonia and alveolar injury were absent when treated with other mAbs. In addition, P1-A10, P1-S3, CN9+CN10, and CN8+CN14 showed minimal inflammation with absent macrophages, lymphocytes, plasma cells, and fibrin. Although many of the treatment animals had residual alveolar hemorrhage and edema, only one of the treatment animals had mild interstitial pneumonia (C4). In summary, our mAbs were able to increase survival for the infected mice with P1-A10 by 100% survival rate. Additionally, mAb treatment, except for C4, exhibited significant protection against lung pathology.

**Figure 6:**
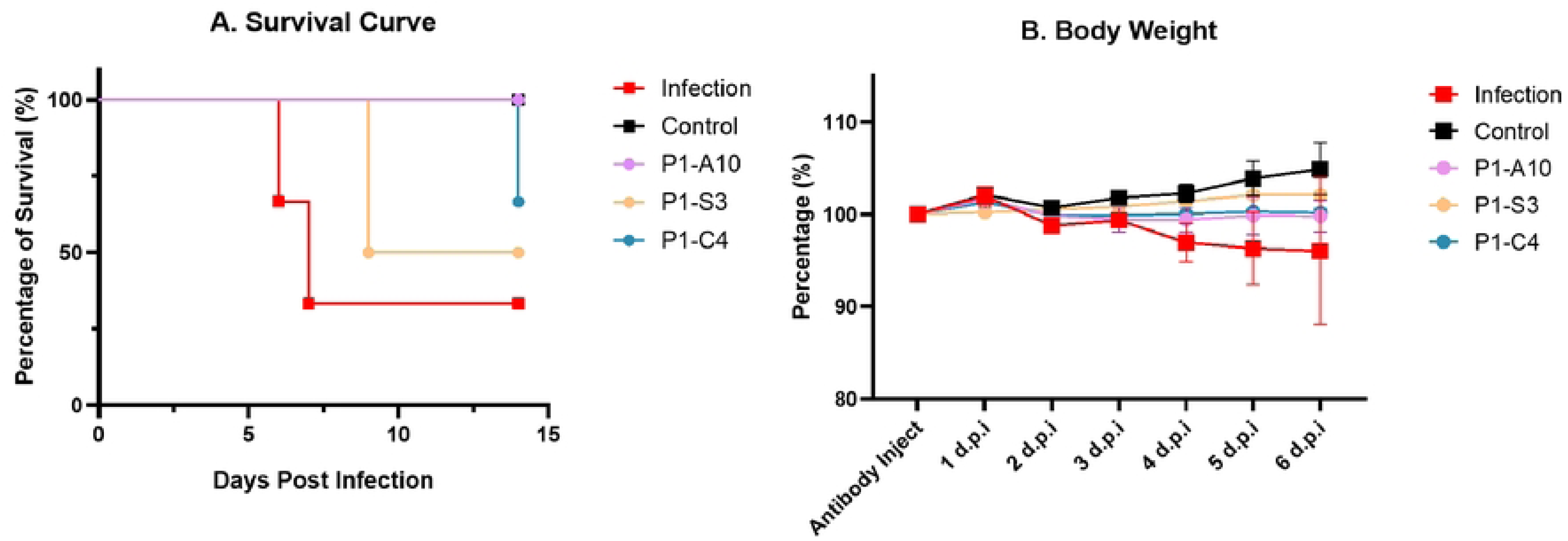
COVID-19 patient-generated mAbs protect mice from SARS-CoV-2 infection. **A**) Survival monitoring of mice 14 days after prophylactic application of SARS-CoV-2 neutralizing mAb and viral challenge. Infection: n=3 on days 1-5, n=2 on day 6; n=3 in control and P1-C4 groups, n=2 in P1-A10 and P1-S3 groups. **B**) Monitoring of mouse body weight 6 days after prophylactic application of SARS-CoV-2 neutralizing mAbs and viral challenge, mean ± SEM for each group of animals. Day 6 (infection versus P1-A10: p=0.135, infection versus P1-S3, p=0.01, infection versus P1-C4, p=0.05) using two-tailed Mann-Whitney test.

## Discussion

The study successfully generated and characterized SARS-CoV-2-specific antibodies from the memory B cells of convalescent COVID-19 patients, particularly targeting the spike protein and RBD. Using SCAN technology and FACS, live memory B cells secreting IgG antibodies were isolated, revealing significant diversity in antibody specificities shaped by individual viral exposure history. The study produced 17 monoclonal antibodies encompassing various heavy and light variable genes, with only a few common between patients. In vitro assays demonstrated varying degrees of inhibition against wild-type and Omicron strains, highlighting discrepancies between ACE2 competition and actual neutralization capacity. Binding kinetics, analyzed using bio-layer interferometry, showed that binding affinity alone does not fully reflect inhibitory potential. Structural analysis through *in silico* docking revealed unique binding motifs, with notable differences in neutralization abilities based on epitope targeting. In vitro reactivity assays confirmed that certain antibodies have consistent inhibition across various concentrations, emphasizing the importance of RBD binding in selecting high-affinity antibodies. Lastly, in vivo experiments using a mouse model demonstrated significant therapeutic potential, with most antibodies reducing inflammation and lung pathology. Overall, this study underscores the complexity of the humoral immune response to SARS-CoV-2 and the need for a multifaceted approach to developing effective therapeutic antibodies and vaccines capable of adapting to the evolving virus.

Patient 1, who was infected with SARS-CoV-2 WA1, produced monoclonal antibodies that inhibited RBD binding to the ACE2 receptor, as validated through the competitive ELISA assay. The D614G mutation did not significantly affect the inhibition ability of these antibodies, suggesting that their binding occurs through interactions with other epitopes that may not include D614G, as most B-cell epitopes range in length from 5 to more than 20 amino acids. Patient 2, infected by SARS-CoV-2 WA-1 and unidentified variants, produced antibodies with broad immunity to original strains and emerging variants. The memory B cells from this patient reacted against N501Y, D614G, and Wuhan or Delta S1 protein in single or cross-reactive manners, as demonstrated by flow cytometry and SCAN sort results. The subsequent pseudovirus neutralization experiment validated their ability to neutralize the Omicron variant, even though it was not included in the sorting criteria. This finding suggests that selecting against a combination of relative epitopes with identical mutations could generate cross-reactive antibodies for other variants. Interestingly, the selected mAbs from patient 1 that exhibited the highest inhibition ability in competitive ELISA increased pseudovirus uptake by cells, ranging from 100% to 200%. Considering the possible inefficiency of original mAbs against emerging variants containing multiple mutation sites, this might be due to ineffective binding, changing the surface alignment of the spike protein, and revealing new antigenic epitopes that facilitate viral uptake.

Compared to the public database, including published and patented mAbs, our lab-produced mAbs have unique characteristics that could dictate their function, as binding to different structural domains could elicit different protective responses. Many effective monoclonal antibodies target the RBD of the S1 subunit, neutralizing the virus by blocking its ability to bind to ACE2. The S2 subunit is more conserved across different coronaviruses compared to the S1 subunit, particularly the RBD; therefore, antibodies targeting the S2 subunit exhibited broader neutralizing activity against multiple variants of SARS-CoV-2^20–22^. Studies have shown that the ultrapotent human antibodies bound to RBM blocked S attachment to ACE2, showing neutralization potencies, and protected hamsters against SARS-CoV-2 challenge^10,23^. Our structural modeling indicated that P1-A10 binds to the RBM in the RBD. This binding could contribute to the challenged animal’s survival, a decrease in lung pathology, and inflammation. P1-S3 binds NTD, which is part of the art of the S1 and is located adjacent to the RBD. Although the NTD does not directly interact with the ACE2, it can influence the overall conformation and stability of the S protein^24^. By binding to the NTD, these antibodies can induce conformational changes that may impede the RBD from effectively binding to the ACE2, thus neutralizing the virus^25^. The NTD might play a role in the conformational changes required for viral fusion with the host cell membrane. Therefore, antibodies that bind to the NTD could disrupt these processes, preventing the virus from entering the host cell. P1-S3 was not as effective as P1-A10 in challenged animals’ survival; however, it exhibited the same protection in the lung pathology and inflammation as P1-A10. Therefore, the two isolated mAbs showed consistency with previous studies in regard to their effectiveness in protecting against SARS-CoV-2 infection.

In conclusion, the study highlights the intricate and multifaceted nature of the humoral immune response to SARS-CoV-2, showcasing the diversity and specificity of antibodies generated from convalescent COVID-19 patients. By employing cutting-edge technologies, a range of monoclonal antibodies was successfully isolated and characterized, with distinct binding affinities and neutralization capacities against various SARS-CoV-2 strains, including challenging variants like Omicron. The study revealed that while some antibodies effectively block viral entry by targeting the RBD and preventing ACE2 binding, others bind to conserved regions such as the S2 or influence the conformation of the S protein through the NTD, thereby inhibiting viral fusion and entry mechanisms.

## Author Contributions

YS performed the isolation and characterization of the monoclonal antibodies using *in vitro* assays, analyzed the data, and wrote the first draft of the manuscript. JT examined the lung pathology. AV assisted with the isolation of monoclonal antibodies. AT conducted the *in vivo* therapeutic effect of the antibodies. CQN conceptualized, analyzed, and assisted with the manuscript writing.

## Declaration of Interest

All authors have no competing interests in the subject of the study.

## Data Availability

All relevant data are within the manuscript and its Supporting Information files.

## Acknowledgements

This research was funded by the National Institutes of Health (NIH) and the National Institute of Dental and Craniofacial Research (NIDCR) (DE028544, DE028544-02S1, PI-Nguyen).

## Notes

### Competing Interest Statement

The authors have declared no competing interest.

### Funding Statement

The author(s) received no specific funding for this work.

### Author Declarations

This study received approval from the University of Florida's Institutional Review Board, with the reference number IRB202001475.

